# Modeling SARS-CoV-2 RNA Degradation in Small and Large Sewersheds

**DOI:** 10.1101/2021.09.17.21263708

**Authors:** Camille McCall, Zheng N. Fang, Dongfeng Li, Andrew J. Czubai, Andrew Juan, Zachary LaTurner, Katherine Ensor, Loren Hopkins, Phil Bedient, Lauren B. Stadler

**Affiliations:** Department of Civil and Environmental Engineering, Rice University, Houston, TX, 77005; Department of Civil Engineering, The University of Texas at Arlington, Arlington, TX, 76019; Department of Statistics, Rice University, Houston, TX, 77005; Houston Health Department, 8000 N. Stadium Dr., Houston, TX, 77054

**Keywords:** COVID-19, Wastewater-based epidemiology, Coronaviruses, Decay, Travel time

## Abstract

Wastewater-based epidemiology has been at the forefront of the COVID-19 pandemic, yet little is known about losses of SARS-CoV-2 in sewer networks. Here, we used advanced sewershed modeling software to simulate SARS-CoV-2 RNA loss in sewersheds across Houston, TX under various temperatures and decay rates. Moreover, a novel metric, population times travel time (PT), was proposed to identify localities with a greater likelihood of undetected COVID-19 outbreaks and to aid in the placement of upstream samplers. Findings suggest that travel time has a greater influence on viral loss across the sewershed as compared to temperature. SARS-CoV-2 viral loss at median travel times was approximately two times greater in 20°C wastewater between the small sewershed, Chocolate Bayou, and the larger sewershed, 69th Street. Lastly, placement of upstream samplers according to the PT metric can provide a more representative snapshot of disease incidence in large sewersheds. This study helps to elucidate discrepancies between SARS-CoV-2 viral load in wastewater and clinical incidence of COVID-19. Incorporating travel time and SARS-CoV-2 decay can improve wastewater surveillance efforts.

## 1. Introduction

Municipal wastewater treatment plants collect untreated wastewater from communities ranging from hundreds to millions of inhabitants per day within a given sewershed. This wastewater can be scrutinized to obtain critical insights into biological and chemical markers that are reflective of community health within the serviced population, an approach known as wastewater-based epidemiology (WBE).

In WBE, untreated wastewater is considered analogous to a population-wide urine and stool sample. This representative sample can be used to evaluate community health and the prevalence of certain diseases by directly measuring markers of concern. Viral monitoring in wastewater has gained much attention considering that viruses do not replicate independent of a host cell and are persistent in the environment. Several viral pathogens including hepatitis A virus, hepatitis E virus, norovirus, sapovirus, astrovirus, and poliovirus have been monitored in wastewater for community health tracking (Bisseux et al., 2018; Brouwer et al., 2018; Kamel et al., 2010; Kokkinos et al., 2011; la Rosa et al., 2014).

Recently, WBE has been recognized as a promising tool for tracking SARS-CoV-2, the causative agent of Coronavirus Disease 2019 (COVID-19). SARS-CoV-2 is an enveloped positive-sense RNA virus belonging to the *Coronaviridae* family. Although the primary transmission route of SARS-CoV-2 is via respiratory droplets, evidence of fecal shedding of SARS-CoV-2 in infected individuals has led to the global attention of WBE in the ongoing fight against COVID-19 (Parasa et al., 2020). Several studies have highlighted the potential for viral signals to precede clinical cases and capture the extent of asymptomatic individuals that are not reported in health care facilities (la Rosa et al., 2021; Randazzo et al., 2020; Stadler et al., 2020; Wu et al., 2020). Generally, evidence supports the utility of WBE as a public health and environmental tracking tool for viral disease outbreaks. Still, in some cases discrepancies exist between viral signal in wastewater and disease prevalence, specifically with SARS-CoV-2 (Ahmed et al., 2021).

Viral measurements from wastewater alone may not be sufficient for disease tracking. Considerations such as the environmental matrix, sampling regimen, sewer collection system, viral stability, and disease characteristics are critical aspects to establishing correlations between viral signal and disease incidence in the community (Xagoraraki, 2020). Among these critical considerations is the stability of the virus and its genetic material in the sewershed. Microbial degradation plays a significant role in determining what proportion of RNA shed in feces gets captured at the outfall of a wastewater treatment plant (WWTP). To date, few studies have investigated RNA degradation of SARS-CoV-2 in wastewater (Ahmed et al., 2020; Bivins et al., 2020; Hart and Halden, 2020a; Weidhaas et al., 2021).

In two of the studies, temperature had a significant influence on variations between first-order decay rates (Ahmed et al., 2021; Weidhaas et al., 2021). Temperature was also found to have a greater impact on RNA degradation than the sample matrix (Ahmed et al., 2020). Despite the agreement on the importance of temperature between the two studies, Weidhaas et al., 2021 obtained a significantly higher decay constant (4.32 day^-1^) at 35°C than Ahmed et al., 2020 (0.24 day^-1^) at 37°C for similar gene targets. This indicates that there are other factors that have a notable influence on RNA degradation such as sample preparation or wastewater composition. A recent study demonstrated that the abundance of the SARS-CoV-2 N1 marker is associated with total organic carbon and pH (Hong et al., 2021). Furthermore, Bivins et al. 2020 evaluated changes in decay constants when the starting viral titer was low as compared to high titers. Low titers (10^3^) obtained a decay constant of 0.09 day^-1^ at 20°C which was significantly lower than that of the high titer (10^5^) at the same temperature, 0.67 day^-1^.

From these studies it is evident that further work is needed to understand degradation of SARS-CoV-2 under various conditions. Moreover, only one study to date has explored degradation of SARS-CoV-2 in sewer systems using a first-order decay rate derived from a study on the infectivity of various coronaviruses in wastewater after 21 days (Gundy et al., 2009; Hart and Halden, 2020a). The authors found that larger sewersheds further confound the effects of temperature on degradation. Indeed, it is expected that longer travel times will create notable discrepancies between viral concentration and COVID cases in communities. Furthermore, since upstream sampling provides high spatial resolution and is more representative of the sampled population (Hart and Halden, 2020a; Matus et al., 2019), the placement of wastewater samplers in sewersheds remains an ongoing area of interest (Larson et al., 2020; Yeager et al., 2020). Yet, to our knowledge, no quantitative approaches for selecting upstream sampling locations based on minimizing degradation of SARS-CoV-2 signal have been proposed.

Here, we model SARS-CoV-2 RNA loss in sewersheds across Houston that vary in service population and geographic area based on published and experimentally derived first-order decay rates, wastewater temperature, and sewershed travel times. Finally, we propose a novel metric for determining critical locations for placing upstream samplers to improve SARS-CoV-2 monitoring in wastewater.

## 2. Materials and Methods

### 2.1. Study Area and Overview

Houston has 39 sewersheds with a total service area covering approximately 1,451 km^2^ (358,580 acres). Of those, ten sewersheds were selected for this study based on the availability of sewershed hydraulic models provided by Houston Public Works. The location and characteristics of the selected sewersheds are detailed in **Figure 1** and **Table S1**, respectively. Hydraulic modeling was conducted to simulate performance metrics which were then used to compute travel times for each sewershed. Multiple SARS-CoV-2 decay rates based on published and experimental studies were then used with the computed travel times to estimate viral persistence in the sewersheds.

**Figure 1.**
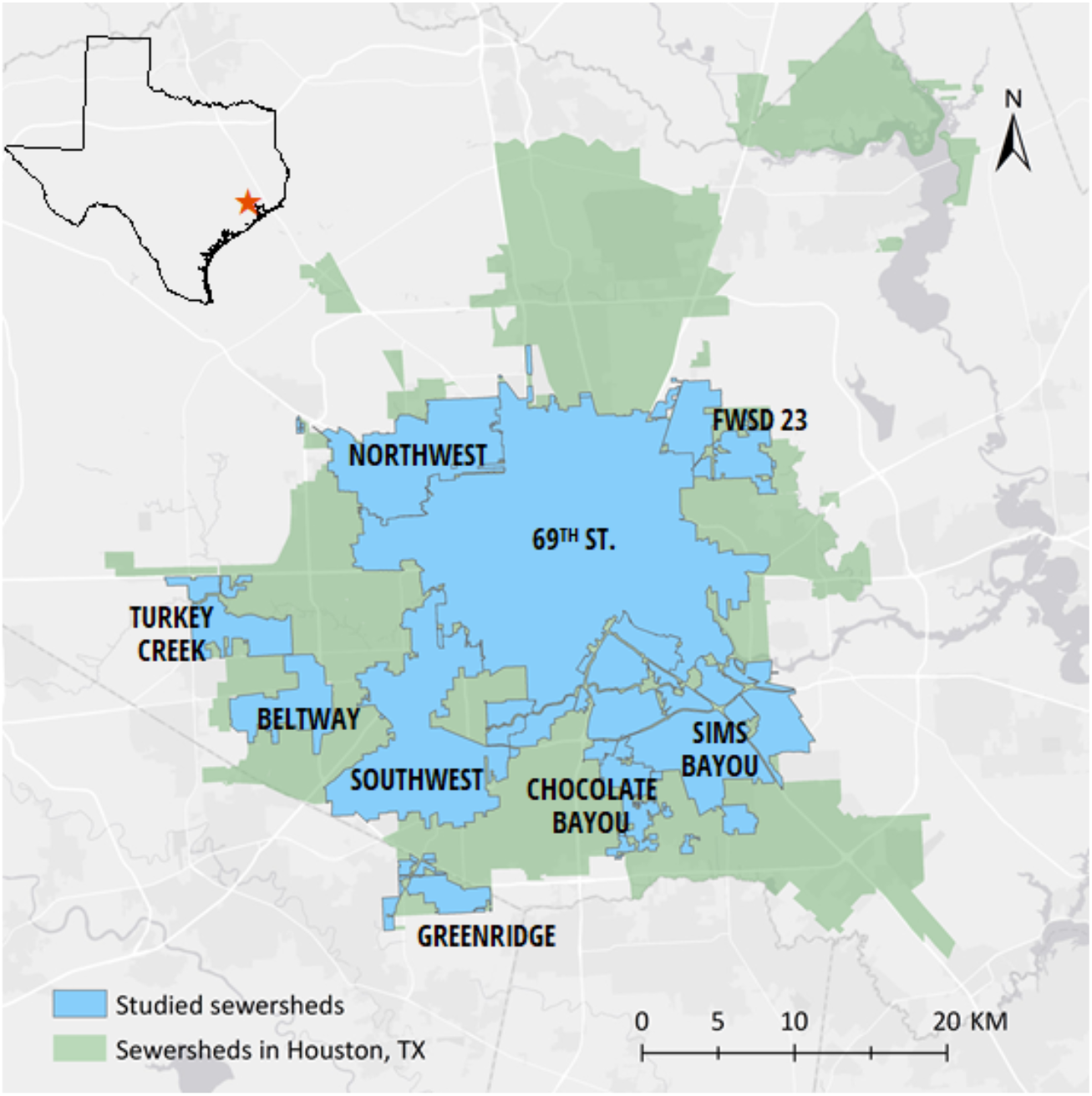
Ten selected wastewater treatment plant service areas (sewersheds) are shown in blue. Sims Bayou has overlapping service areas recognized as Sims Bayou North and South sewersheds. The remainder of Houston’s sewersheds are shown in green.

### 2.2. Sewershed Modeling

Hydraulic modeling of sewersheds in this study was accomplished using the Infoworks ICM software (ICM stands for Integrated Catchment Modeling). Developed by Innovyze®, Infoworks ICM is a hydrodynamic model capable of simulating the hydrology and hydraulics of aboveground surfaces as well as underground drainage networks based on the conservation of mass and momentum. Due to its robustness and versatility, Infoworks ICM has been used by numerous municipalities, like the City of Houston, for stormwater, flood control infrastructure, and sewer management.

In this study, Infoworks ICM models for ten sewersheds were obtained from the City of Houston Public Works department. Each model represents the wastewater network and service areas for a sewershed. Infoworks ICM provides separate solution models for permeable planes, force mains, pressurized pipes or normal gravity flow. An ICM model consists of a network of links and nodes, in which the links represent pipes or conduits, and the nodes represent manholes or other control structures (e.g., outfall or WWTP). Additionally, the model allows for either one or multiple outfall locations. Based on the connectivity of the nodes and links, service areas that drain to any particular node could be further separated into individual subcatchments. The gradient or slope of the link is calculated using the provided starting and ending invert elevations. ICM divides each conduit into a number of discrete computational points and regularly-spaced segments with intervals that are 20 times the pipe diameter. Flow, velocity, and other performance metrics are computed in each segment. Inflow can be added to certain nodes as point sources, but in this study, diurnal curves in the form of wastewater profiles with hourly time steps were applied at corresponding subcatchments to represent wet and dry conditions. With specified information of population and per capita flow, the wastewater profile can be developed from a calibrated model and is designed to mimic dry weather flow as typically seen during flow monitoring.

### 2.3. Computing Travel Time

While various performance metrics such as head, flow, velocity, volume, and water depth are computed by ICM, individual travel times for subcatchments are not. In a typical wastewater system, the conduits are connected to a common outfall, which usually represents the local WWTP. All model networks must have at least one common outfall, but networks are allowed to have multiple outfalls which could represent wet-weather overflows, emergency bypasses, or pump stations to a different wastewater system. Because there is not always a single common outfall, a structured query language (SQL) script was developed to allow users to specify a terminal point. Having terminal points enables travel time to be computed from any subcatchment to the specified points. The user determines the number of iterations for the query, the minimum assumed velocity, and the specific simulation time that the query would run. The query then uses the trace tool, which selects all upstream conduits, nodes, and subcatchments to a specified point and iterates the simulated results to determine a corresponding travel time based on the cumulative conduit length travelled and velocity at the given point. In the case where multiple flow paths exist, the query assumes that wastewater would always travel on the shortest path, therefore computing the shortest travel time from the point of entry to the terminal point. Lastly, the computed travel times for the entire model network were then exported into GIS for further analysis.

### 2.4. Identifying Decay Rate Studies

A literature search was conducted in October 2020 and again in February 2021 using Web of Science and Google Scholar databases to identify studies with first-order degradation rates for SARS-CoV-2 RNA. SARS-CoV-2 was used as a keyword paired with one or more of the following: wastewater, degradation, decay, sewershed, persistence, fate, and survivability. The criteria for inclusion were (1) peer-viewed journal articles (excluded reviews, metadata, pre-prints, editorial material), (2) a focus on SARS-CoV-2 in untreated wastewater samples or simulated untreated wastewater, (3) includes at least one original, experimentally determined decay rate for SARS-CoV-2 RNA.

### 2.5. Decay of SARS-CoV-2 RNA in Sewage

Along with experimentally determined decay rates of SARS-CoV-2 from published literature, decay rates were also generated in our lab. To determine decay rates for SARS-CoV-2 RNA, roughly 1 gallon of wastewater influent was collected from a 24-hour composite sampler and transported on ice to Houston Public Works central processing laboratory. Approximately 500 ml of wastewater was collected in triplicate, stored in Nalgene bottles, and transported on ice to Rice University. The 500 ml bottles were weighed to accurately determine the volume of wastewater in each bottle. Next, each sample was poured into a sterilized 1 L Erlenmeyer flask containing a stir bar. Flasks were loosely capped with aluminum foil to prevent evaporation and placed on a stir plate at the lowest possible setting to maintain a heterogenous mixture.

A 50 ml sample was immediately collected from each flask and concentrated via an HA filtration method. HA filters were stored at -80°C. Wastewater from each flask was collected, concentrated, and stored via this method every 24 hours for the next 6 days. All wastewater samples were incubated at room temperature (∼20°C) in a Biosafety cabinet. After the 6 days, all stored samples were simultaneously extracted using the Qiagen Allprep Powerviral DNA/RNA kit (Qiagen).

SARS-CoV-2 N1 and N2 gene targets were quantified in wastewater extracts using previously described primers and probes (LaTurner et al., 2021). A duplex reverse transcriptase digital droplet PCR (RT-ddPCR) was carried out using the One-Step RT-ddPCR Advanced kit for probes (Bio-Rad) on a QX200 AutoDG Droplet Digital PCR System (Bio-Rad) according the manufacturer’s recommendations. Ten microliters of RNA extract, no template control, or positive control was transferred to a 12 *μ*l reaction mix containing final concentrations of 900 nmol of each primer and 250 nmol for each probe. All reactions were performed in triplicate with thermocycling conditions detailed here (LaTurner et al., 2021). Samples were then read on a QX200 Droplet Reader (Bio-Rad) and analyzed using the QuantaSoft v1.7.4 software. The limit of quantification was previously determined as 0.767 gene copies/*μ*l. Further details of sample concentration, extraction, and quantification can be found in (LaTurner et al., 2021). A linear regression analysis was performed in R (R Core Team 2020) to determine the decay rates for each target. Concentrations of SARS-CoV-2 were log-transformed to satisfy the assumptions of normality according to a visual inspection of the quantile-quantile (Q-Q) plots.

### 2.6. Estimation of SARS-CoV-2 RNA Signal Loss in Sewersheds Based on First-order Decay

To the best of our knowledge, all experimentally-derived published decay rates for SARS-CoV-2 were included in this study. A temperature of 20°C was used to compare the influence of each decay rate on the proportion of viral load loss in select sewersheds across studies. The following formula is an approximation of the Arrhenius equation used to determine the dependence of first-order rates on temperature (Behradek, 1930; Blaustein et al., 2013; Hart and Halden, 2020a):

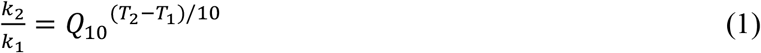

where *Q*_10_ is the temperature coefficient, *k*_1_and *k*_2_ are the lower and upper decay rate constants, respectively, and *T*_1_ and *T*_2_ are the temperatures in Celsius for the upper and lower rate constants, respectively. The temperature coefficient *Q*_10_ is the factor by which a rate changes given a ten degree increase in temperature and is usually between 2 and 3 for biological systems (Behradek, 1930; Reyes et al., 2008).

Eq (1) was used to estimate the decay rate at 20°C for the Weidhaas et al., 2021 study, using the decay rates measured at 10 and 35°C. The temperature-dependent linear regression equation reported by the authors was used to determine the decay rate of SARS-CoV-2 RNA at 20°C for Ahmed et al., 2020.

The degradation of SARS-CoV-2 in the sewershed over time is expected to follow exponential decay as expressed in eq (2) where C(t) is the concentration of SARS-CoV-2 after time *t, C*_0_ is the initial concentration of SARS-CoV-2 released in the wastewater, and *k* is the first order decay rate.

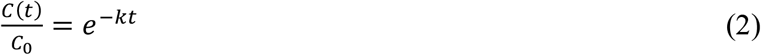

Assuming an initial viral RNA proportion of 1 or 100%, eq (2) was substituted into eq (3) to estimate the proportion of SARS-CoV-2 RNA loss (L) eq (4) or remaining (R) eq (5) at a given time within the sewershed.

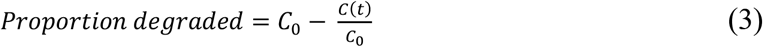

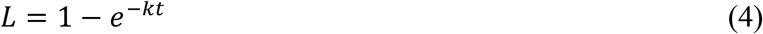

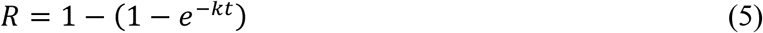

The half-life 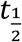 and *t*_90_ (the time for viral load to decrease by one log unit) for each decay rate *k* were obtained from the published work or derived from the following formulas, respectively:

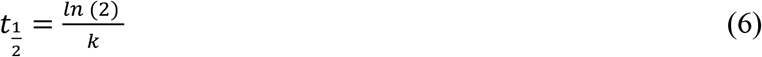

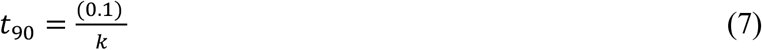

### 2.7. PT Metric for Identifying Hotspots

Aside from using travel time isochrones to determine the spatial distribution of viral persistence, they could also be used in conjunction with population density information to help identify potential viral hotspot areas. In this study, a normalized population times travel time (PT) metric is introduced to identify critical locations for placing upstream samplers. Normalized PT maps for the 69th Street sewershed were generated by multiplying population density information (P) in each sub-sewershed area by their corresponding travel times (T), and then normalized by the maximum PT value computed for the 69th Street and Chocolate Bayou sewersheds. The resulting normalized PT maps have values that range from 0 (0%) to 1 (100%). Areas with low PT values indicate a low likelihood of an undetected outbreak, due to low population density, short travel times, or both. Conversely, areas with high PT values imply a higher likelihood of undetected outbreaks. This is especially true for areas with the highest PT values (i.e., at or close to 100%), signifying that those areas have both high population density and long travel times.

## 3. Results and Discussion

### 3.1. Impact of Weather Conditions on Wastewater Travel Time in Sewersheds

To assess the impact of wet weather on travel times, the ICM model was used to compute travel times under wet and dry conditions for each sewershed (**Figure 2**). Travel times under dry weather conditions were generally higher than wet conditions. The 69th Street and Chocolate Bayou sewersheds were used for further analysis due to the differences in characteristics, and because they represented the sewersheds with the largest and smallest service areas studied, respectively. Median dry weather travel times for 69th Street and Chocolate Bayou were 523 min (s.d. = 217.58 min) and 220 min (s.d.= 152.12 min) with a comparable maximum dry weather travel time of 1207 mins and 1123 mins, respectively (**Figure 3**).

**Figure 2.**
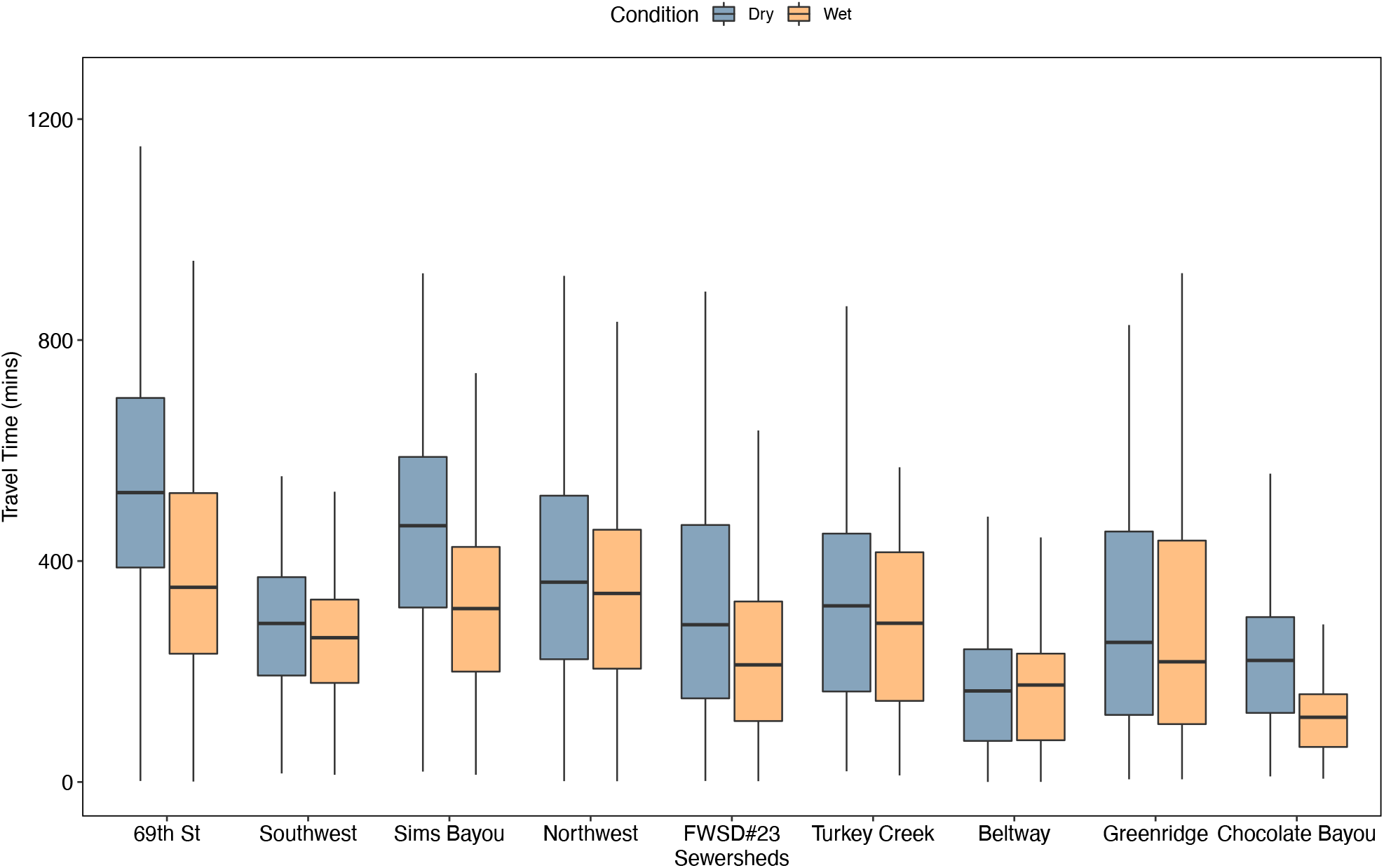
Boxplot of travel times for select sewersheds under dry and wet weather conditions. Horizontal lines represent the median travel time. Lower and upper whiskers represent the 25^th^ and 75^th^ percentiles, respectively.

**Figure 3.**
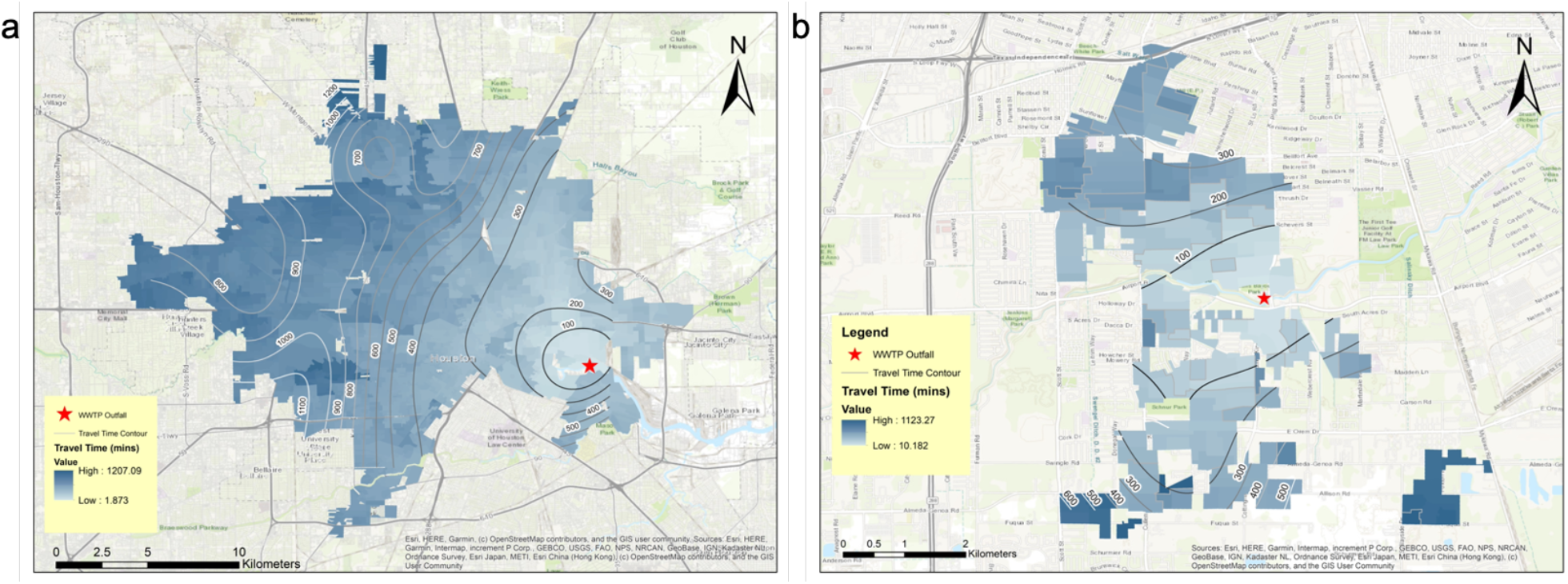
Heat map displaying travel time for 69th Street (a) and Chocolate Bayou (b). Numbers indicate travel time isochrones in minutes from the WWTP outfall (indicated with a red star).

### 3.2. Impact of Decay Rate and Temperature on SARS-CoV-2 Loss in Transit to Wastewater Treatment Plant

We determined the decay rates of SARS-CoV-2 N1 and N2 at 20°C using wastewater collected from a sewershed in Houston to compare values using a Houston-specific wastewater to previously published decay rates. After the fourth day of incubation, unclear concentration dynamics occurred wherein the concentration of all targets slightly increased. Due to uncertainty in the cause of this behavior, only the first few days were considered in the regression analysis. Degradation of N1 and N2 showed similar behavior with decay rates of 0.84 day^-1^ and 0.82 day^-1^, respectively. **Table S2** displays the linear regression parameters for each gene. Our experimentally-determined decay rates were within the range of published rates (**Table 1**). Decay rates listed in **Table 1** were used to evaluate the impact of decay rate and temperature on virus loss in Houston sewersheds.

**Table 1.** Summary of reported decay rates, half-lives, and t_90s_ (the time for viral concentration to decrease by one log unit) for SARS-CoV-2 RNA targets in various studies at 20°C.

| Duration of Experiment (day) | Duration of Experiment (hr) | Target | Decay Rate, $k$ ($\text{day}^{-1}$) | Half Life, $t_{1/2}$ (day) | $t_{90}$ (day) | Comments | Reference |
| --- | --- | --- | --- | --- | --- | --- | --- |
| 33 | 792 | N1 | 0.15 | 4.78 | 15.88 |  | Ahmed et al. 2020 |
| 1 | 24 | N1, N2 | 1.75 | 0.40 | 1.31 |  | Weidhaas et al. 2020 |
| 6 | 144 | N1 | 0.84 | 0.83 | 2.74 |  | In lab |
| 7 | 168 | E | 0.67 | 0.99 | 3.30 | High titer | Bivins et al. 2020 |

There were significant differences in how the decay rates in **Table 1** were determined, which may explain the wide range of reported rates. (Weidhaas et al., 2021) obtained the fastest decay rates compared to studies considered with a rate of (1.75 day^-1^) at 20°C. The authors measured SARS-CoV-2 RNA in wastewater samples obtained from two different treatment plants immediately after collection. These initial concentrations were then compared to concentrations measured in replicate samples incubated at 4, 10, and 35°C for 1 to 22 hours. Ahmed et al., 2020 spiked SARS-CoV-2-negative wastewater samples with RNA extracted from gamma-irradiated SARS-CoV-2 hCoV-19/Australia/VIC01/2020 isolate and incubated them at 4, 15, 25, and 37°C over the course of 33 days in which RNA concentrations from those samples were measures every few days.

Decay rates from Bivins et al., 2020 were the most congruent with results from our lab except under low titer conditions (starting concentration of 10^3^) (data not shown). The decay rate under low titer conditions was significantly slower than all other reported decay rates listed here (Bivins et al., 2020). Here, the authors inoculated non-sterile wastewater with a SARS-CoV-2 isolate from a clinical patient diagnosed with COVID-19 at low titer (10^3^) and high titer (10^5^) concentrations. SARS-COV-2 RNA was extracted and quantified in 20°C inactivated wastewater samples over the course of 7 days. The decay rate associated with the high titer SARS-CoV-2 concentration was selected from the Bivins et al., 2020 study because it was more representative of concentrations previously measured in Houston sewersheds. Notably, the fastest reported decay rates from Weidhaas et al., 2021 and our own experiments were determined in samples that were not spiked with virus. This may have been due to the form of the virus in wastewater samples, which is likely a mixture of intact, protected (enveloped and/or intact capsid) virus, and degraded unprotected viral RNA. Degraded, unprotected viral RNA will degrade much faster than intact, protected virus (Wurtzer et al., 2021).

Given the ability to estimate decay rates at various temperatures for values obtained from Ahmed et al., 2020 and Weidhaas et al., 2021, and because they represented the lowest and highest decay rates reported to date, respectively, these studies were used to evaluate the effect of wastewater temperature (20-30°C) on the fraction of RNA loss over time. As expected, viral RNA loss increases with increasing travel time. Moreover, travel time has a greater influence on virus loss as compared to temperature within the range of travel times estimates for the ten sewersheds considered in this study (**Figure 4**). However, it is important to note that the impact of temperature on virus loss increases with increasing travel times as displayed in **Figure 4**. For example, the difference in the percent of virus loss between 20 and 30°C is 0.6% and 9.8% for Ahmed et al., 2020 and Weidhaas et al., 2021, respectively after a travel of 120 min compared to 5.2% and 16% at 1200 min.

**Figure 4.**
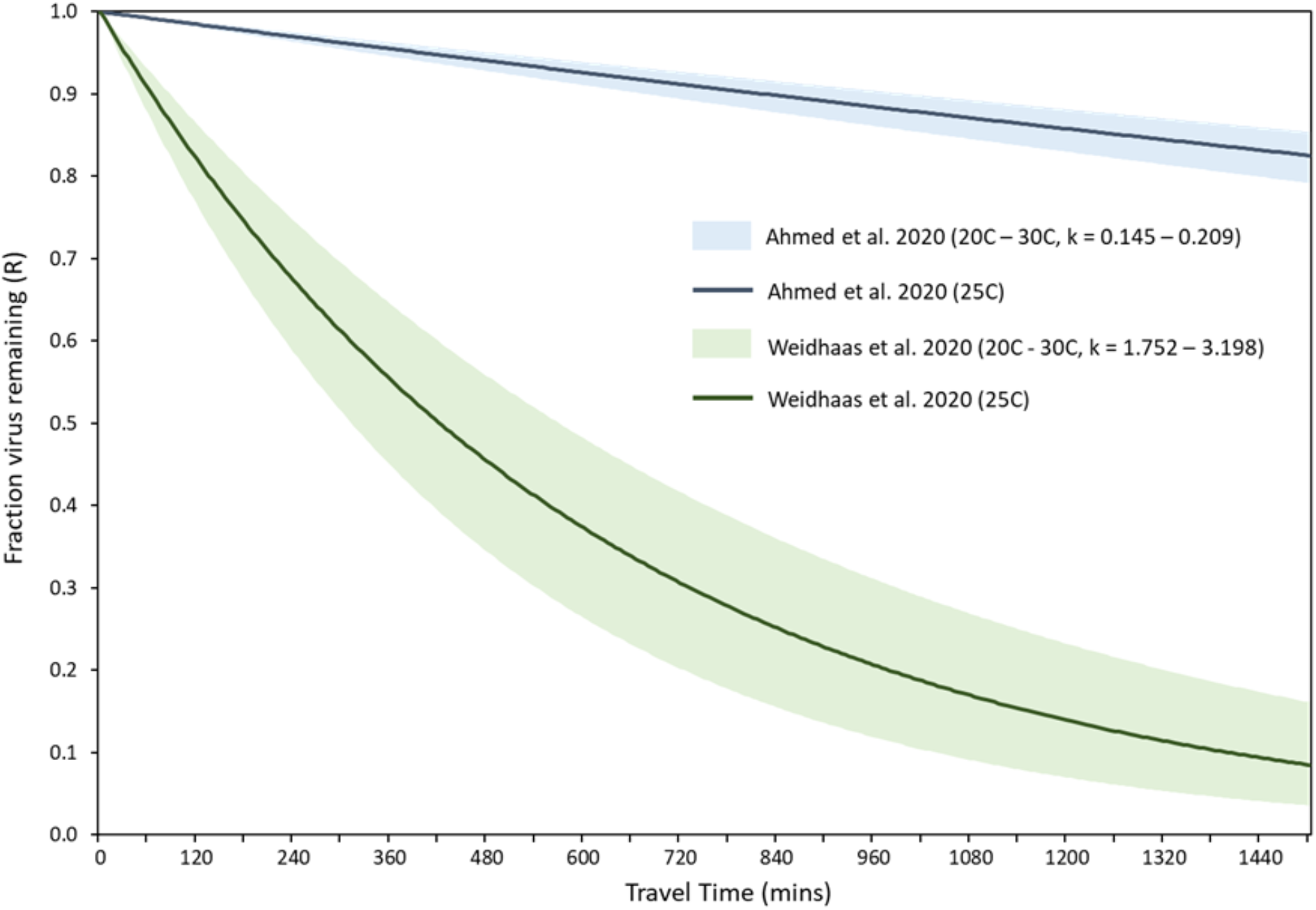
Effects of temperature, decay rate, and travel time on virus loss.

Similar findings were reported in a recent study that assessed SARS-CoV-2 RNA in sewersheds in Tempe, Arizona under varying wastewater temperatures (Hart and Halden, 2020a). The authors concluded that under high temperature conditions in large sewersheds, viral concentration at outfalls may be less representative of disease incidence as compared to colder temperatures. Wastewater temperatures can fluctuate by as much as 27°C depending on geographical region and seasonal changes (Hart and Halden, 2020b). Therefore, careful consideration of wastewater temperatures can be used to improve disease prevalence estimations and explain discrepancies in correlations between the number of disease cases and virus concentrations.

The percent of SARS-CoV-2 RNA loss in wastewater traveling from a given geographical location within the 69th Street and Chocolate Bayou sewersheds to their corresponding outfalls at the WWTPs was determined using travel time and decay rates from **Table 1**. As expected, 69th Street showed greater variability in viral loss across the sewershed as compared to Chocolate Bayou. Viral losses at a median travel time of 523 min for 69th Street were 5.13, 21.60, 26.29, and 47.08% for the 0.145, 0.670, 0.840, and 1.752 day^-1^ decay rates, respectively. Chocolate Bayou obtained median percent losses of 2.19, 9.73, 12.04, and 23.48% at a median travel time of 220 min (**Figure 5**). Taking into consideration the decay rate obtained from our study, approximately a 25% reduction in viral signal is estimated in the 69th Street sewershed compared to a 12% reduction for Chocolate Bayou.

**Figure 5:**
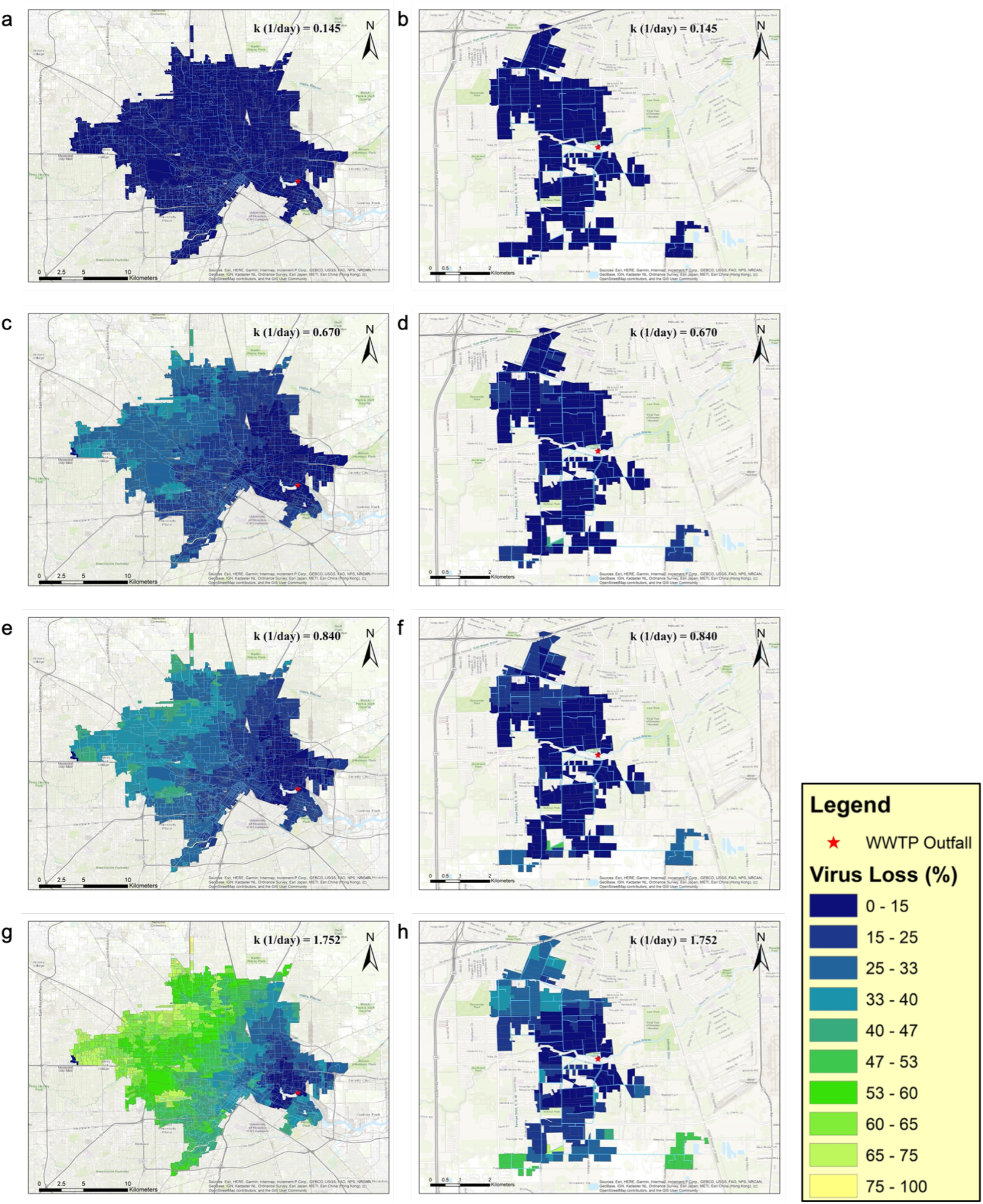
Geographical heat maps of SARS-CoV-2 RNA loss for 69th Street (a, c, e, g) and Chocolate Bayou (b, d, f, h) under dry weather conditions and decay rates obtained or estimated from studies listed in Table 1.

Decay rates of 0.840, and 1.752 day^-1^ resulted in a virus loss of approximately ≥ 50% when considering travel times between 1190 and 570 min, respectively. Travel time range between 0-1123 min for the Chocolate Bayou sewershed, thus all regions maintained less than a 50% reduction in virus loss for a decay rate of 0.840 day^-1^. Despite a greater reduction of viral loss in both sewersheds when considering a virus degradation rate of 1.752 day^-1^, the fraction of the 69th Street sewershed with ≥ 50% viral loss is 49.84% as compared to 2.98% for Chocolate Bayou. Consequently, virus decay is more critical in the 69th Street sewershed due to the number of remote regions.

### 3.3. Population vs. Travel Time Metric

To account for virus decay across large sewersheds we propose PT, a novel metric used to facilitate the placement of upstream samplers and minimize travel times throughout sewersheds. Upstream sampling refers to sampling within the sewer system from locations such as manholes, as compared to sampling at the influent of a WWTP (downstream sampling). There are practical constraints that dictate the selection of upstream sites, namely available resources, accessibility, and safety. Additionally, site selection based on population density and travel time should be considered to further increase the impact of upstream sampling on WBE outcomes.

The PT metric identifies areas that are at high risk of a wide-spread COVID-19 outbreaks due to population density and the outbreak is less likely to be fully captured in WBE due to prolonged travel times. To evaluate the efficacy of this metric, we estimated PT values for all regions within the 69th Street sewershed. **Figure 6a** illustrates a PT heatmap in the case of downstream sampling only. Hotspots were identified in the northwest-central region of the sewershed.

**Figure 6.**
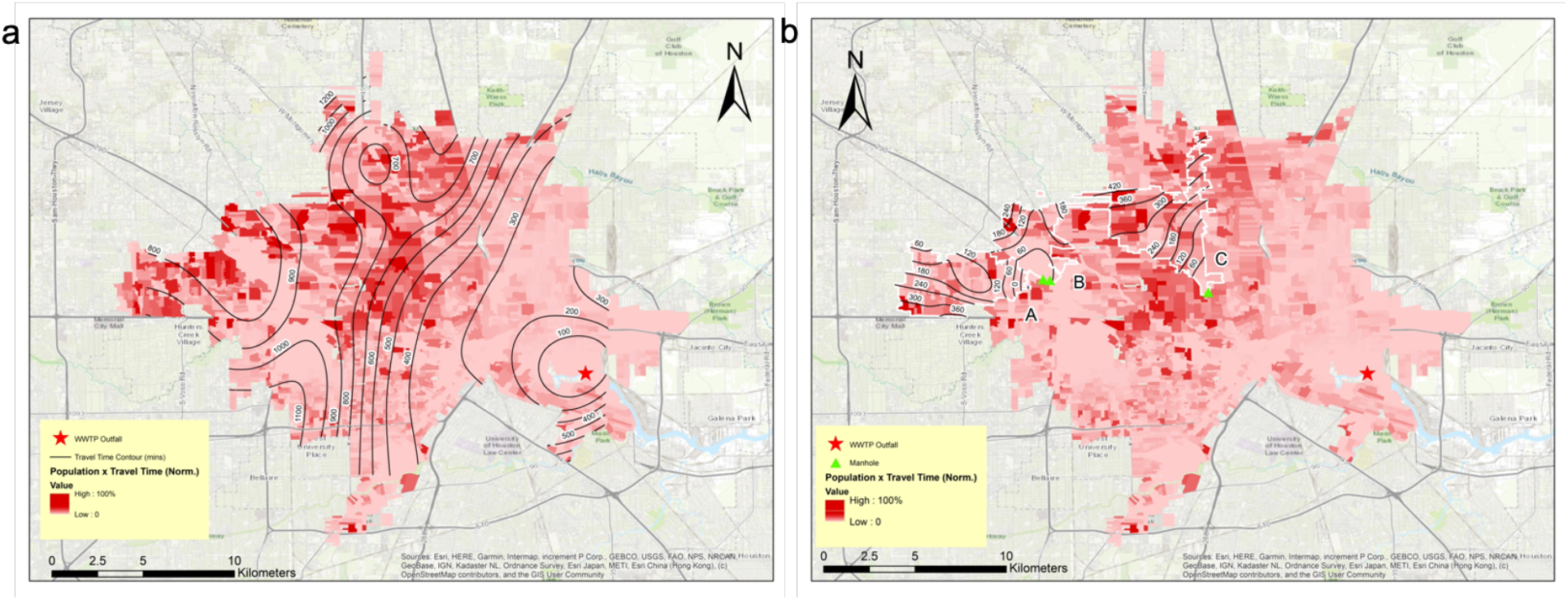
(a) Display of hotspots in dark red according to normalized PT values and (b) placement of samplers and adjusted travel times in sub-catchment zones A, B, and C in the 69th Street sewershed.

A second simulation was carried out with three upstream samplers hypothetically placed in hotspots (higher PT values), located in the northwest-central region, that were expected to reduce travel times in the sub-sewershed areas and minimize PT throughout the sewershed (**Figure 6b**). Placement of samplers decreased the median travel time in zones A, B, and C as indicated in **Figure 6b** from 865, 840, and 869 min to 154, 129, and 313 min, respectively. Results here indicate that implementation of upstream samplers according to the PT metric can significantly reduce the number of hotspots in large sewersheds.

## 4. Limitations and Implications

The scope of this study does not directly take into account other factors that could influence virus degradation such as wastewater composition and microbial predation, which could all further explain or impact conclusions presented here. Since average travel times are approximated from the ICM models, they may not strictly reflect transit of wastewater in the sewersheds, particularly in remote locations and during varying diurnal cycles. Still, findings here indicate that in-sewer decay may be an important factor to consider in WBE and when designing sampling campaigns. The modeling approach used here can be adopted in various localities to improve environmental surveillance efforts and public health responses to local and global viral disease outbreaks.

## 5. Conclusion

- Travel time has a greater impact on viral loss across sewersheds as compared to temperature.
- Degradation in large sewersheds can be up to two times higher than small sewersheds.
- The use of the PT metric for placement of upstream samplers can reduce travel times in large sewersheds by more than 60%.
- Considering travel time may improve WBE for COVID-19 and account for discrepancies in correlations between cases and viral concentrations.

## Data Availability

Data available upon request.

## Acknowledgment

A special thanks to Houston Public Works for providing ICM models for selected sewersheds and additional sewershed characteristics where needed. We’d also like to thank the Houston Health Department, and specifically Courtney Hundley, Rebecca Schneider, and Lilian Mojica, for providing aggregate travel times for positive COVID-19 cases as needed. We thank Prashant Kalvapalle for his contribution to the wastewater-based epidemiology method development along with the Lauren Stadler and Anthony Maresso lab groups at Rice University and Baylor College of Medicine for their assistance with wastewater sample collection, processing, and analysis.

## References

Ahmed, W., Angel, N., Edson, J., Bibby, K., Bivins, A., O’Brien, J.W., Choi, P.M., Kitajima, M., Simpson, S.L., Li, J., Tscharke, B., Verhagen, R., Smith, W.J.M., Zaugg, J., Dierens, L., Hugenholtz, P., Thomas, K. v., Mueller, J.F., 2020. First confirmed detection of SARS-CoV-2 in untreated wastewater in Australia: A proof of concept for the wastewater surveillance of COVID-19 in the community. Science of the Total Environment. https://doi.org/10.1016/j.scitotenv.2020.138764

Ahmed, W., Tscharke, B., Bertsch, P.M., Bibby, K., Bivins, A., Choi, P., Clarke, L., Dwyer, J., Edson, J., Nguyen, T.M.H., O’Brien, J.W., Simpson, S.L., Sherman, P., Thomas, K. v., Verhagen, R., Zaugg, J., Mueller, J.F., 2021. SARS-CoV-2 RNA monitoring in wastewater as a potential early warning system for COVID-19 transmission in the community: A temporal case study. Science of the Total Environment 761. https://doi.org/10.1016/j.scitotenv.2020.144216

Behradek, 1930. Temperature Coefficients in Biology. Biological Reviews 5, 30–59.

Bisseux, M., Colombet, J., Mirand, A., Roque-Afonso, A.M., Abravanel, F., Izopet, J., Archimbaud, C., Peigue-Lafeuille, H., Debroas, D., Bailly, J.L., Henquell, C., 2018. Monitoring human enteric viruses in wastewater and relevance to infections encountered in the clinical setting: A one-year experiment in central France, 2014 to 2015. Eurosurveillance 23, 1–11. https://doi.org/10.2807/1560-7917.ES.2018.23.7.17-00237

Bivins, A., Greaves, J., Fischer, R., Yinda, K.C., Ahmed, W., Kitajima, M., Munster, V.J., Bibby, K., 2020. Persistence of SARS-CoV-2 in Water and Wastewater. Environmental Science and Technology Letters 7, 937–942. https://doi.org/10.1021/acs.estlett.0c00730

Blaustein, R.A., Pachepsky, Y., Hill, R.L., Shelton, D.R., Whelan, G., 2013. Escherichia coli survival in waters: Temperature dependence. Water Research 47, 569–578. https://doi.org/10.1016/j.watres.2012.10.027

Brouwer, A.F., Eisenberg, J.N.S., Pomeroy, C.D., Shulman, L.M., Hindiyeh, M., Manor, Y., Grotto, I., Koopman, J.S., Eisenberg, M.C., 2018. Epidemiology of the silent polio outbreak in Rahat, Israel, based on modeling of environmental surveillance data. Proceedings of the National Academy of Sciences of the United States of America 115, E10625–E10633. https://doi.org/10.1073/pnas.1808798115

Gundy, P.M., Gerba, C.P., Pepper, I.L., 2009. Survival of Coronaviruses in Water and Wastewater. Food and Environmental Virology 1, 10–14. https://doi.org/10.1007/s12560-008-9001-6

Hart, O.E., Halden, R.U., 2020a. Computational analysis of SARS-CoV-2/COVID-19 surveillance by wastewater-based epidemiology locally and globally: Feasibility, economy, opportunities and challenges. Science of the Total Environment 730. https://doi.org/10.1016/j.scitotenv.2020.138875

Hart, O.E., Halden, R.U., 2020b. Modeling wastewater temperature and attenuation of sewage-borne biomarkers globally. Water Research 172. https://doi.org/10.1016/j.watres.2020.115473

Hong, P.Y., Rachmadi, A.T., Mantilla-Calderon, D., Alkahtani, M., Bashawri, Y.M., al Qarni, H., O’Reilly, K.M., Zhou, J., 2021. Estimating the minimum number of SARS-CoV-2 infected cases needed to detect viral RNA in wastewater: To what extent of the outbreak can surveillance of wastewater tell us? Environmental Research 195. https://doi.org/10.1016/j.envres.2021.110748

Kamel, A.H., Ali, M.A., El-Nady, H.G., Aho, S., Pothier, P., Belliot, G., 2010. Evidence of the co-circulation of enteric viruses in sewage and in the population of Greater Cairo. Journal of Applied Microbiology. https://doi.org/10.1111/j.1365-2672.2009.04562.x

Kokkinos, P., Ziros, P., Meri, D., Filippidou, S., Kolla, S., Galanis, A., Vantarakis, A., 2011. Environmental surveillance. An additional/alternative approach for virological surveillance in Greece? International Journal of Environmental Research and Public Health 8, 1914–1922. https://doi.org/10.3390/ijerph8061914

la Rosa, G., della Libera, S., Iaconelli, M., Ciccaglione, A.R., Bruni, R., Taffon, S., Equestre, M., Alfonsi, V., Rizzo, C., Tosti, M.E., Chironna, M., Romanò, L., Zanetti, A.R., Muscillo, M., 2014. Surveillance of hepatitis A virus in urban sewages and comparison with cases notified in the course of an outbreak, Italy 2013. BMC Infectious Diseases 14, 1–11. https://doi.org/10.1186/1471-2334-14-419

la Rosa, G., Mancini, P., Bonanno Ferraro, G., Veneri, C., Iaconelli, M., Bonadonna, L., Lucentini, L., Suffredini, E., 2021. SARS-CoV-2 has been circulating in northern Italy since December 2019: Evidence from environmental monitoring. Science of the Total Environment 750. https://doi.org/10.1016/j.scitotenv.2020.141711

Larson, R.C., Berman, O., Nourinejad, M., 2020. Sampling manholes to home in on SARS-CoV-2 infections. PLoS ONE 15. https://doi.org/10.1371/journal.pone.0240007

LaTurner, Z.W., Zong, D.M., Kalvapalle, P., Gamas, K.R., Terwilliger, A., Crosby, T., Ali, P., Avadhanula, V., Santos, H.H., Weesner, K., Hopkins, L., Piedra, P.A., Maresso, A.W., Stadler, L.B., 2021. Evaluating recovery, cost, and throughput of different concentration methods for SARS-CoV-2 wastewater-based epidemiology. Water Research 197. https://doi.org/10.1016/j.watres.2021.117043

Matus, M., Duvallet, C., Soule, M.K., Kearney, S.M., Endo, N., Ghaeli, N., Brito, I., Ratti, C., Kujawinski, E.B., Alm, E.J., 2019. 24-hour multi-omics analysis of residential sewage reflects human activity and informs public health. bioRxiv. https://doi.org/10.1101/728022

Parasa, S., Desai, M., Thoguluva Chandrasekar, V., Patel, H.K., Kennedy, K.F., Roesch, T., Spadaccini, M., Colombo, M., Gabbiadini, R., Artifon, E.L.A., Repici, A., Sharma, P., 2020. Prevalence of Gastrointestinal Symptoms and Fecal Viral Shedding in Patients with Coronavirus Disease 2019: A Systematic Review and Meta-analysis. JAMA Network Open. https://doi.org/10.1001/jamanetworkopen.2020.11335

R Core Team (2020). R: A language and environment for statistical computing. R Foundation for Statistical Computing, Vienna, Austria. https://www.R-project.org/.

Randazzo, W., Truchado, P., Cuevas-Ferrando, E., Simón, P., Allende, A., Sánchez, G., 2020. SARS-CoV-2 RNA in wastewater anticipated COVID-19 occurrence in a low prevalence area. Water Research 181. https://doi.org/10.1016/j.watres.2020.115942

Reyes, B.A., Pendergast, J.S., Yamazaki, S., 2008. Mammalian Peripheral Circadian Oscillators Are Temperature Compensated NIH Public Access, J Biol Rhythms.

Stadler, L.B., Ensor, K.B., Clark, J.R., Kalvapalle, P., LaTurner, Z.W., Mojica, L., Terwilliger, A., Zhuo, Y., Ali, P., Avadhanula, V., Bertolusso, R., Crosby, T., Hernandez, H., Hollstein, M., Weesner, K., Zong, D.M., Persse, D., Piedra, P.A., Maresso, A.W., Hopkins, L., 2020. Wastewater Analysis of SARS-CoV-2 as a Predictive Metric of Positivity Rate for a Major Metropolis. medRxiv. https://doi.org/10.1101/2020.11.04.20226191

Weidhaas, J., Aanderud, Z.T., Roper, D.K., VanDerslice, J., Gaddis, E.B., Ostermiller, J., Hoffman, K., Jamal, R., Heck, P., Zhang, Y., Torgersen, K., Laan, J. vander, LaCross, N., 2021. Correlation of SARS-CoV-2 RNA in wastewater with COVID-19 disease burden in sewersheds. Science of the Total Environment 775. https://doi.org/10.1016/j.scitotenv.2021.145790

Wu, F., Zhang, J., Xiao, A., Gu, X., Lee, W.L., Armas, F., Kauffman, K., Hanage, W., Matus, M., Ghaeli, N., Endo, N., Duvallet, C., Poyet, M., Moniz, K., Washburne, A.D., Erickson, T.B., Chai, P.R., Thompson, J., Alm, E.J., 2020. SARS-CoV-2 Titers in Wastewater Are Higher than Expected from Clinically Confirmed Cases. mSystems 5. https://doi.org/10.1128/msystems.00614-20

Wurtzer, S., Waldman, P., Ferrier-Rembert, A., Frenois-Veyrat, G., Mouchel, J.M., Boni, M., Maday, Y., Marechal, V., Moulin, L., 2021. Several forms of SARS-CoV-2 RNA can be detected in wastewaters: Implication for wastewater-based epidemiology and risk assessment. Water Research 198, 117183. https://doi.org/10.1016/j.watres.2021.117183

Xagoraraki, I., 2020. Can We Predict Viral Outbreaks Using Wastewater Surveillance? Journal of Environmental Engineering 146, 01820003. https://doi.org/10.1061/(asce)ee.1943-7870.0001831

Yeager, R.A., Holm, R.H., Saurabh, K., Fuqua, J.L., Talley, D., Bhatnagar, A., Smith, T.R., 2020. Wastewater sample site selection to estimate geographically-resolved community prevalence of COVID-19: A research protocol. medRxiv. https://doi.org/10.1101/2020.08.23.20180224

