## Supplemental Data for "Modeling SARS-CoV-2 RNA Degradation in Small and Large Sewersheds"

Mailing address:

6100 Main Street, MS-519

Houston, TX 77005

**Abstract**

Wastewater-based epidemiology has been at the forefront of the COVID-19 pandemic, yet little is known about losses of SARS-CoV-2 in sewer networks. Here, we used advanced sewershed modeling software to simulate SARS-CoV-2 RNA loss in sewersheds across Houston, TX under various temperatures and decay rates. Moreover, a novel metric, population times travel time (PT), was proposed to identify localities with a greater likelihood of undetected COVID-19 outbreaks and to aid in the placement of upstream samplers. Findings suggest that travel time has a greater influence on viral loss across the sewershed as compared to temperature. SARS-CoV-2 viral loss at median travel times was approximately two times greater in 20°C wastewater between the small sewershed, Chocolate Bayou, and the larger sewershed, 69th Street. Lastly, placement of upstream samplers according to the PT metric can provide a more representative snapshot of disease incidence in large sewersheds. This study helps to elucidate discrepancies between SARS-CoV-2 viral load in wastewater and clinical incidence of COVID-19. Incorporating travel time and SARS-CoV-2 decay can improve wastewater surveillance efforts.

**Keywords:** COVID-19, Wastewater-based epidemiology, Coronaviruses, Decay, Travel time

### Supporting Information

**Table S1.** Characteristics of the ten sewersheds included in this study. Flow rates represent the average daily flow rate under dry conditions for the respective sewershed.

| Sewersheds | 69th Steet | Southwest | Sims Bayou | Northwest | FWSD#23 | Turkey Creek | Beltway | Greenridge | Chocolate Bayou |
|---|---|---|---|---|---|---|---|---|---|
| Service Area ($km^2$) | 274.00 | 98.50 | 96.20 | 58.10 | 26.70 | 22.60 | 22.50 | 17.00 | 15.80 |
| Pipeline Length (km) | 620.00 | 218.78 | 198.00 | 125.00 | 55.00 | 48.00 | 43.00 | 37.00 | 62.00 |
| Service Population | 478,984 | 272,566 | 19,143 | 72,500 | 6,595 | 5,588 | 54,864 | 4,177 | 4,225 |
| Flow rate (liters/day) | 3.22E+08 | 1.09E+08 | 6.91E+07 | 3.32E+07 | 1.17E+07 | 2.17E+07 | 1.40E+07 | 1.23E+07 | 1.05E+07 |

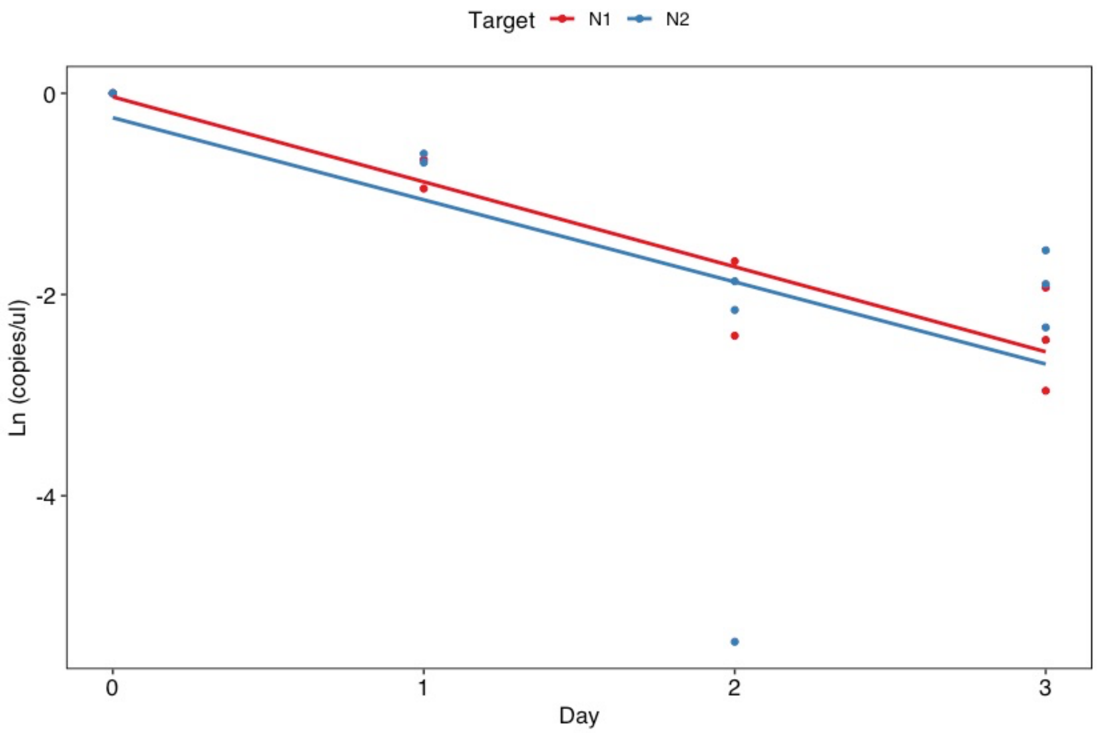


**Figure S1.** Linear regression lines for first-order decay of SARS-CoV-2 RNA for N1 and N2 gene targets.

**Table S2.** Summary of linear regression model for first-order decay of SARS-CoV-2 N1 and N2 gene targets.

| Target | k | SE | $r^2$ | P |
|---|---|---|---|---|
| N1 | -0.84 | 0.10 | 0.90 | 0.00 |
| N2 | -0.82 | 0.34 | 0.39 | 0.04 |
